# *On the cause and consequences of coinfection:*A general mechanistic framework of within-host parasite competition

**DOI:** 10.1101/2024.06.28.24309686

**Authors:** Ashwini Ramesh, Spencer R Hall

**Author notes:** Co-author.

## Abstract

Coinfections pose serious threats to health and exacerbate parasite burden. If coinfection is detrimental, then what within-host factors facilitate it? Equally importantly, what hinders it, say via exclusion or priority effects? Such interactions ought to stem from their within-host environment (‘niche’), i.e., resources that parasites steal from hosts and immune cells that kill them. Yet, despite two decades of empirical focus on within-host infection dynamics, we lack a mechanistic framework to understand why coinfection arises and the diverse range of its’ consequences. Hence, we construct a trait-based niche framework, one that illustrates general principles that govern parasite competition for a resource and apparent competition for immune cells. We show that coinfection requires a competition-resistance tradeoff and that each parasite most impacts the niche factor to which it is most sensitive. These predictions then provide mechanistic interpretation for infection outcomes seen in a variety of extant experiments: Why does nutrient supplementation shift relative frequencies of coinfecting parasites? When and how does sequence of parasite invasion allow only early invading parasites to win? How does intrinsic variation in immune response shape coinfection burden? Together, this mechanistic framework of parasite competition offers new perspectives to better predict within-host infection dynamics and improve individual health.

## INTRODUCTION

Coinfections pose a serious threat to health both at the individual and population scale for hosts. Broadly, coinfection is the successful concurrent infestation of a host with two or more parasite species. Coinfection worsens human health (76% of 2000 publications in a meta-analysis) and exacerbates infection burden (57%; Griffiths *et al* 2011). For instance, bacterial co-infections cause two-fold increase in mortality in COVID patients (Shah *et al* 2023). Similarly, coinfecting hghhhelminth species increased the odds of anaemia five-to-eight-fold among children (Ezeamama *et al* 2005). Coinfection at the individual scale can also influence population-scale disease dynamics (Ezenwa & Jolles 2011). Populations having higher frequency of coinfection experience larger epidemics than those with epidemics of one parasite (Susi *et al* 2015). Yet, despite virulent costs, coinfections are pervasive amongst hosts, including humans, wildlife, and livestoc/ agriculture (Vogels *et al* 2018; Ezenwa *et al* 2010; Halliday, Penczykowski *et al* 2020). Therefore, we need to better understand why and how parasites coinfect their hosts.

Nonetheless, coinfection represents just one of numerous outcomes of concurrent pathogenesis. That fact poses fundamental questions about coinfection. First, if coinfection is indeed detrimental, then what within-host factors facilitate it? Equally importantly, what prevents it? Within-host parasite competition can lead to coinfection (*i.e.,* within-host parasite coexistence), singly infection (through exclusion or priority effects), or clearance (no infection) from hosts (Vogel *et al* 2018). Yet, we lack theoretical predictions to elucidate those outcomes of competition. For instance, irrespective of timing, sequential exposure to some parasites always lead to coinfection, yet to others, only early infecting parasites prevail (Fig. 1A; Clay *et al* 2019; Devevey *et al* 2015). Second, when they coinfect why do some parasites become more abundant than others in relative and absolute senses? For instance, nutrient supply or resources can shift this ‘community structure’ of parasites within hosts favouring one parasite over the other (Fig. 1B; Fellous & Koella 2009, Budischak *et al* 2015). Finally, not all coinfected hosts are equal. Some hosts present higher coinfection burden than others, hinting at variation in immunological resistance (Fig. 1C; Fuess *et al* 2021, Halliday *et al* 2018). How does such variation govern coinfection burden? Presently, these disparate bits about coinfection remain unconnected, lacking a synthetic glue. Here, we seek to catalyse creation of a framework linking genesis of coinfection to its consequences (Lively *et al* 2014; Restif & Graham 2015).

**Figure 1:**
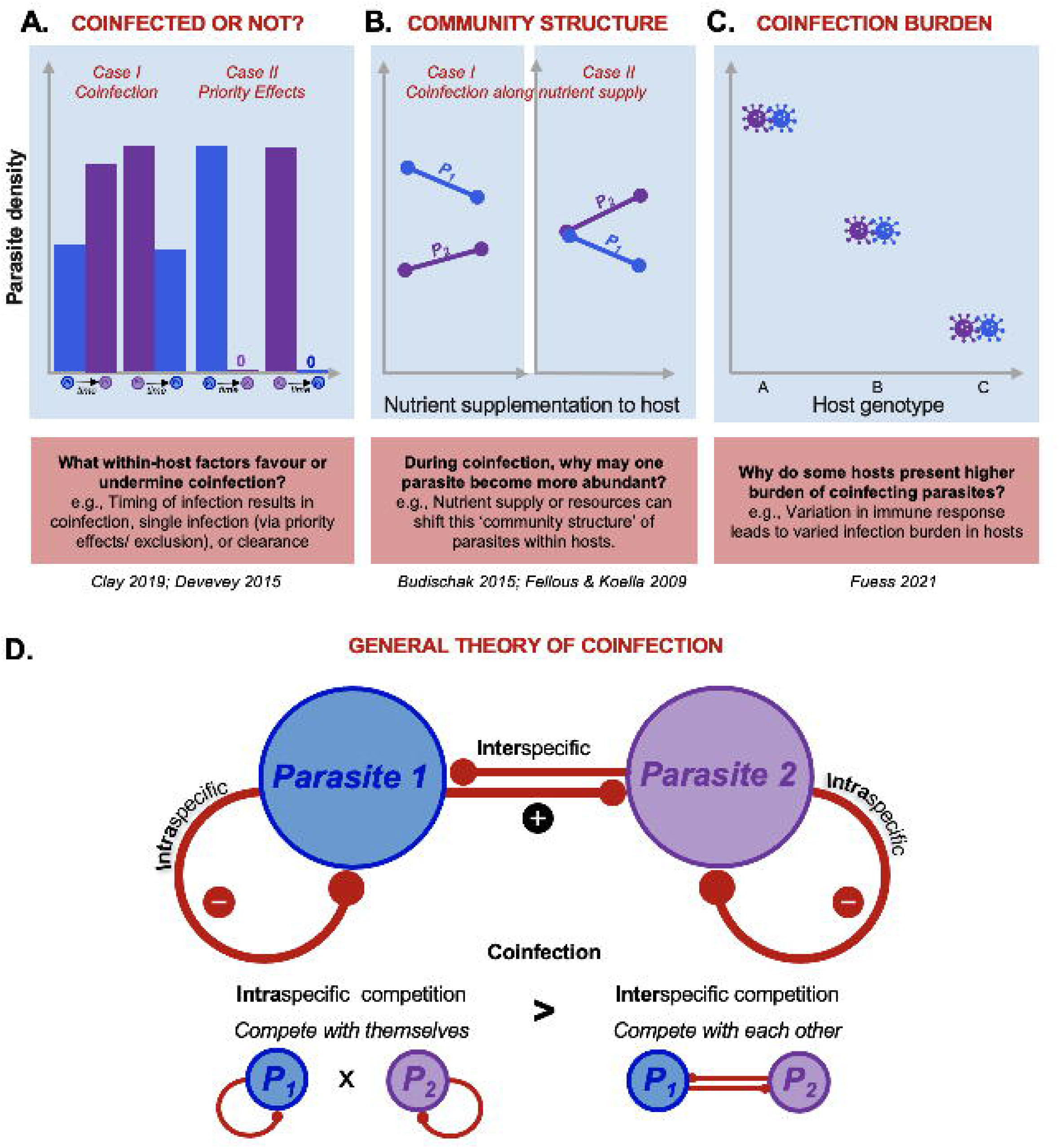
*Unpacking causes and consequences of coinfection:* (A-C) Examples from experiments probing why coinfection arises and the diverse range of its’ consequences, including: (A) *Coinfected or not?* What within-host factors facilitate coinfection and what prevents it? Sequential exposure to parasite species can lead to coinfection irrespective of timing (Case I); in other scenarios, early-infecting parasites prevailed (impeding coinfection; Case II). (B) *Coinfection community structure:* When they coinfect, why do some parasites become more abundant than others? When can nutrient supply or resources increase a parasite’s relative abundance (Case I) and both relative and absolute abundance (Case II)? (C) *Coinfection burden:* Finally, some hosts or host genotypes present higher (co)infection burden than others hinting at differences in immunological resistance. (D) Parasite competition hinges on two main interactions: how (a) species compete with themselves or intra-specific competition (negative intraspecific direct effect, red curve), and how (b) species compete interspecifically (negative interspecific direct effect, red arrows). Generally, coinfection (or within-host coexistence) occurs when intra-exceeds inter-specific competition, and vice-versa for priority effects, at a feasible ‘interior’ equilibrium (Table S2). Here, those outcomes are illustrated with direct effects (in the Lotka-Volterra model). Subsequently, those competitive effects work indirectly through niche factors (like energy or immune cells).

A mechanistic, within-host framework of parasite competition provides a start. To illustrate, we develop and evaluate within-host models of coinfection, synthesizing insights through an ecological lens (following Ramesh & Hall 2023). Broadly, coinfection (*i.e.,* within-host coexistence of parasites) occurs when each parasite species competes more strongly with themselves (intraspecifically) than with each other (interspecifically; Fig. 1D). Those competitive differences could be measured phenomenologically. Even better, they can be quantified from trait-based niche models for parasites competing within hosts (Graham *et al* 2008, Cressler *et al* 2014). In such models, parasites interact within host ‘ecosystems’ for shared resources while facing attack by energetically costly immune cells. Simple ecological rules, then, govern within-host parasite competition. To illustrate, we first borrow from old and new niche models to outline how shared energy (resources) and/or immune cells could govern divergent coinfection outcomes (Fig. 2). Second, using a case study of a two parasite – immune cells – energy model (2PIE) niche model, we link outcomes of infection to competitive abilities, nullclines (niches), feedback, and key traits (Figs. 3-7; Table S1,2). Third, our approach scripts how to link *a priori* predictions to interpretation of experimental outcomes using these types of models (Fig. 8-9). Finally, we offer suggestions for future theory and experiments (Fig. 10, Table 1). Taken together, we lay a general mechanistic framework to understand coinfection using within-host niche-based competition model.

**Figure 2:**
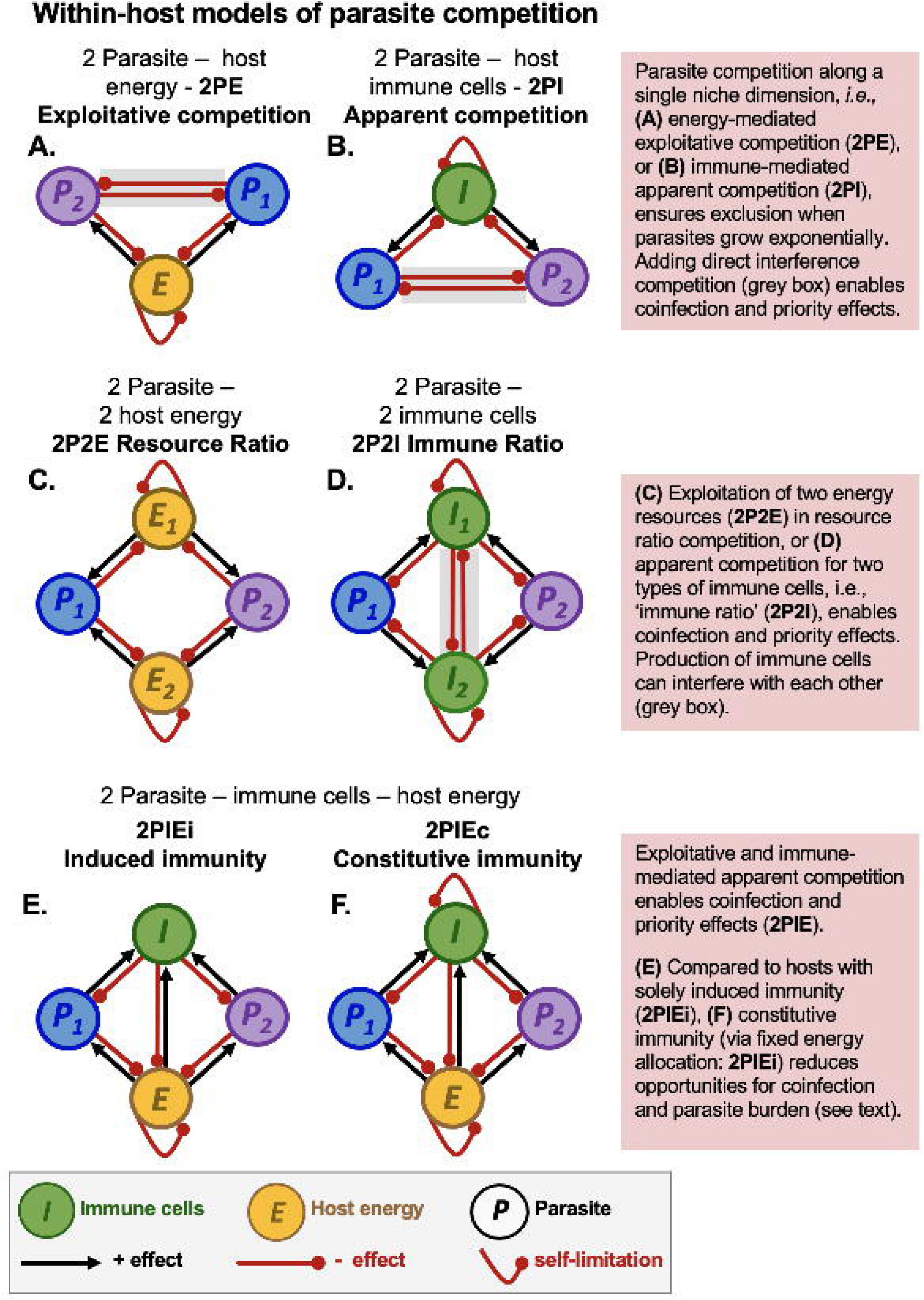
*Models of within-host competition of parasites, with potential infection outcomes*. *Top row, single host energy or immune cells model*: Two parasites can compete indirectly for (A) host energy (exploitative: 2PE) or (B) via immune cells (apparent competition: 2PI). Parasites can also engage directly in interference competition (denoted by *P_1_ – P_2_* interactions: grey shading). *Middle row, two energy or immune models:* Two parasites can compete for (C) two sources of energy or other resources within hosts (2P2E; i.e., like a resource ratio model) or for (D) two different forms of immune cells or classes (2P2I; ‘immune ratio’; mutual inhibition between *I_1_ – I_2_* is possible [grey shading]). *Bottom row, energy and immune models:* Competing parasites can be attacked by shared immune cells and compete for host energy with (E) induced immunity (2PIEi), and (F) with constitutive immunity (2PIEc). Immune cells require host energy for proliferation resulting in additional *E-I* loops beyond those above (B, D). Red (black) arrows: negative (positive) interspecific direct effects, evaluated at positive densities (a feasible interior equilibrium); red curved arrow: self-limitation (negative intraspecific effect).

**Figure 3:**
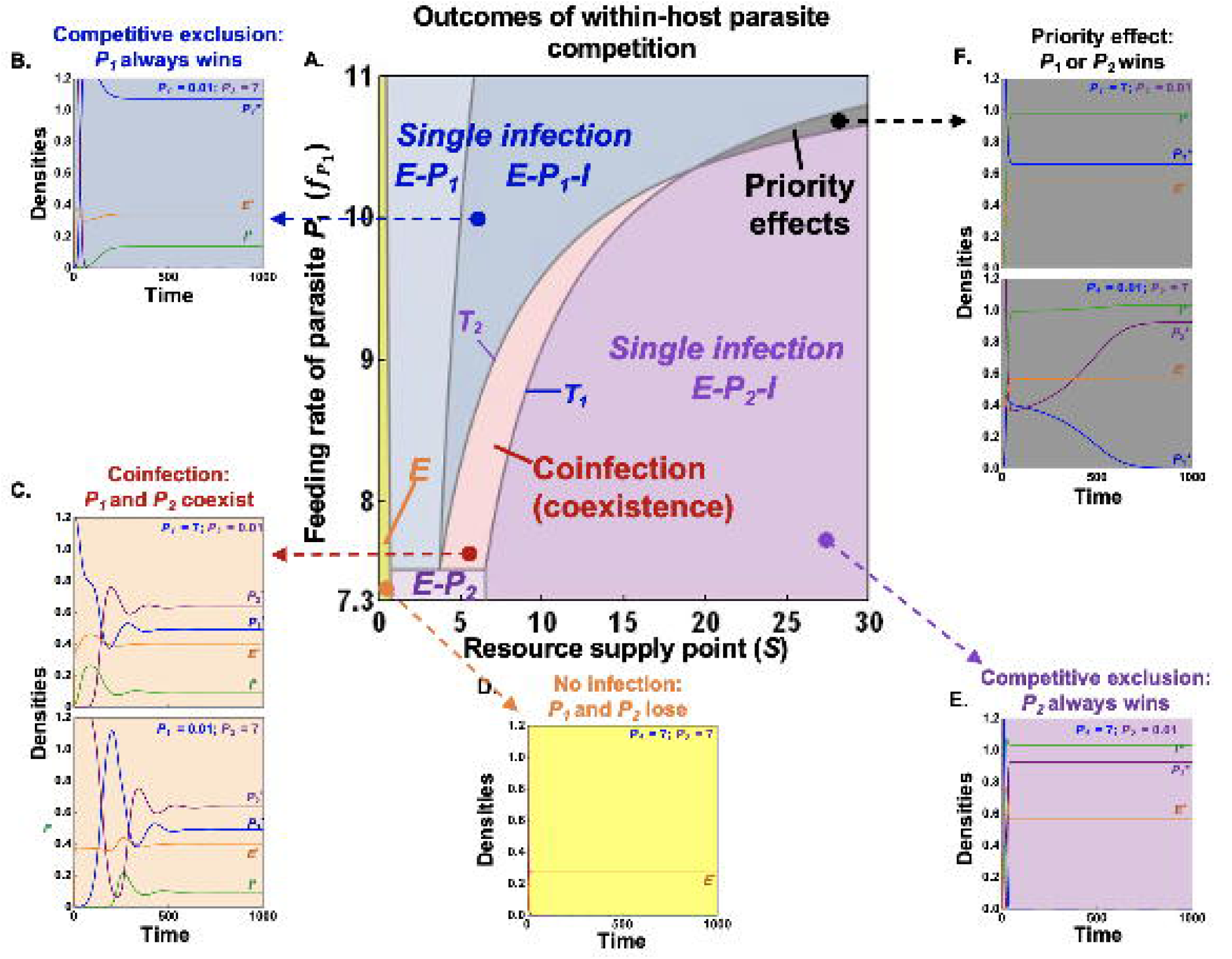
*Outcomes of within-host competition in the 2PIEi model* (A) Outcomes of competition are characterized here in a 2D-bifurcation diagram over gradients of nutrient supply point (*S*) and feeding rate of a parasite, 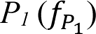. The (a)symmetry in inter- and intra-specific competition involved governs single infection (via competitive exclusion: *P_1_* [blue] or *P_2_* [purple] only), coinfection (orange), priority effects (gray), or no infection (yellow). (B – F) Sample dynamics for one or two starting densities of *P_1_* and *P_2_*.

**Table 1.**
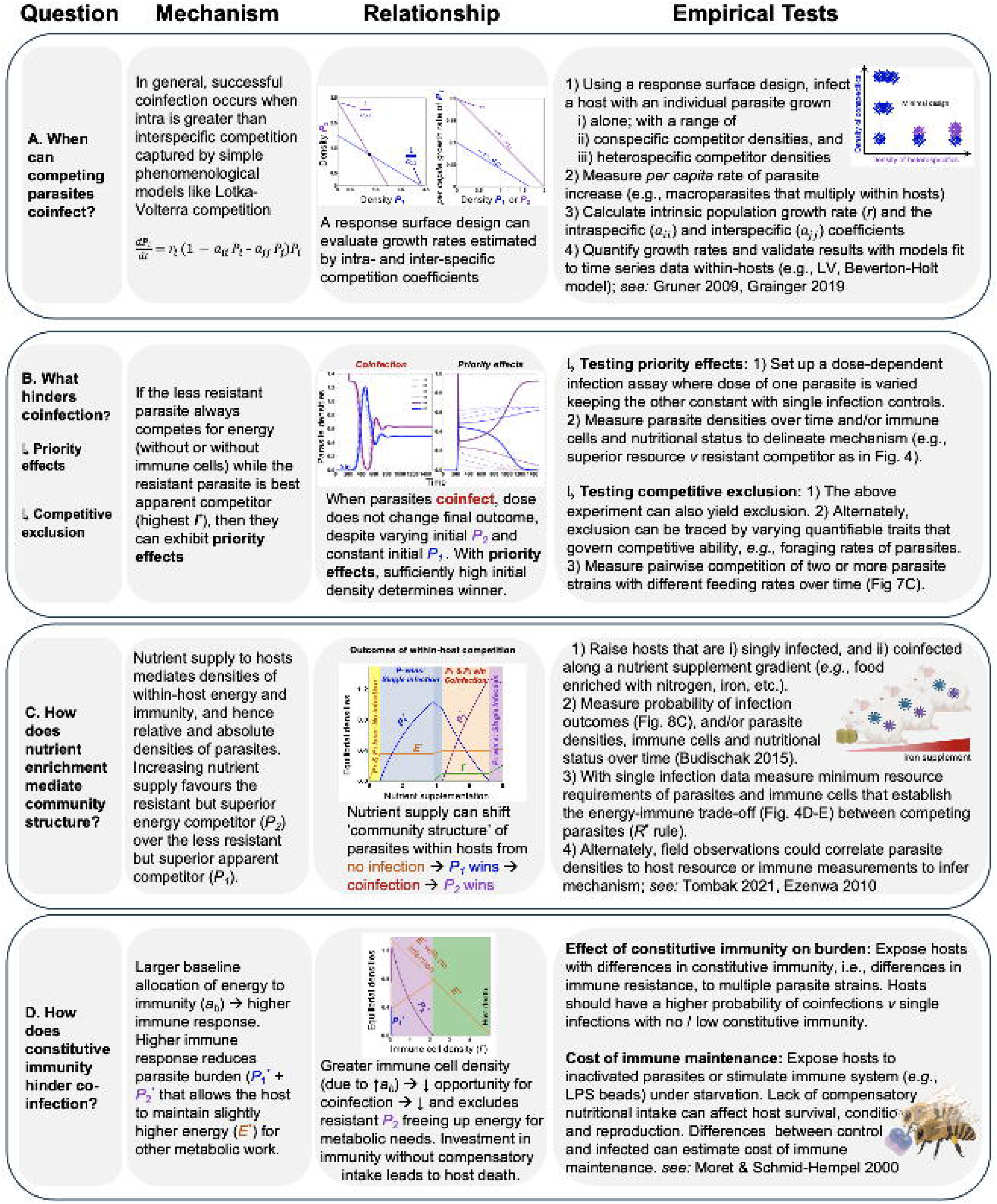
*A guide to future coinfection experiments:* The outcomes of within-host competition can be tested by (A) measuring species’ invasion growth rates via phenomenological models (e.g., Lotka-Volterra), or (B-D) via parameterizing a model mechanistic model like those outlined in Fig. 2 – 7. ‘Questions’ outline fundamental queries surrounding causes and consequences of divergent infection outcomes. ‘Mechanisms’ summarize the theory, and ‘Relationships’ shows equations or correlations that can connect theory to experiments. ‘Empirical Tests’ provides examples of how those relationships can be tested across a range of systems. Figure created using <u>BioRender.com</u>.

### Section I: A phenomenological approach to coinfection

To visualize these interactions, we label the direct effect of species *j* on growth rate of species *i*, yielding (hereafter) interspecific *positive effects* (black arrow) and *negative effects* (red arrow) and intraspecific *self-limitation* (red curve; Fig. 1,2). Then, we can write the two-species Lotka–Volterra competition model in terms of intra (*a_ii_*)- and inter (*a_ij_*)-specific per capita competition coefficients:

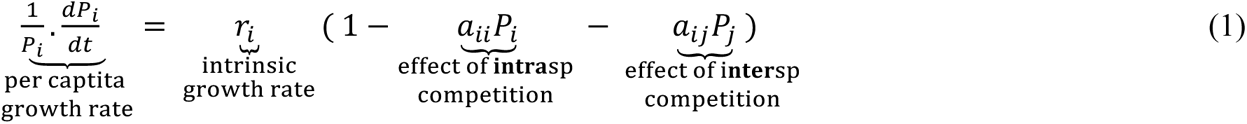

where *P_i_* is the density of parasite species *i*, and *r_i_* is its intrinsic (maximum per capita) growth rate. This model assumes that per capita growth rates (fitness) decrease linearly with density of each species with slopes *a_ii_* and *a_ij_* (Table 1A). Successful coinfection satisfies two conditions. First, each single-species (‘boundary’) carrying capacity (1/*a_ii_*) must be invasible by the other parasite when rare. That condition is met when each species reduces its own growth rate more than that of its competitor (i.e., *a_11_* > *a_21_* and *a_22_* > *a_12_*). Second, joint intraspecific (*a_11_ a_22_*) must exceed joint interspecific competition (*a_12_ a_21_*). When both are met, the feasible coexistence (‘interior’) equilibrium is stable with negative feedback (Table 1A).

Coinfection is hindered in one of two ways. First, strong positive feedback at the interior generates priority effects when joint inter-exceeds intra-specific competition (*a_12_ a_21_* > *a_11_ a_22_*). Priority effects also require mutually uninvasible boundary equilibria and a feasible interior (which are both met when *a_11_* < *a_21_* and *a_22_* < *a_12_*). Here, high enough initial density of parasites determines the winner. Second, competitive exclusion arises with asymmetric competition. Specifically, if *P_2_* cannot invade *P_1_*’s boundary (*a_11_* < *a_21_*) but *P_1_* can invade *P_2_*’s boundary (*a_22_* > *a_12_*), then *P_1_* excludes *P_2_*. Conversely, *P_2_* excludes *P_1_* when *a_11_* > *a*_21_ but *a_22_* < *a_12_*. Hence, mutually or asymmetrically strong interspecific competition can hinder coinfection.

Empirically, estimates of competition coefficients can come from densities and growth rates of parasites. Various methods exist, including a response surface design. In the context of macroparasites, this design involves growing species either alone or at the lowest densities that lead to infection. It also involves growing species with increasing density of intra- and inter-specific competitors. Through this design, *a_ii_* and *a_ij_* can be estimated, then used to predict if species can successfully infect when rare (Freckleton & Watkinson 2000; *reviewed in* Hart *et al* 2018; see Table 1A). However, this approach remains phenomenological, measuring coefficients only in a specific environment. That context-dependence limits predictions of infection outcomes across gradients of traits, nutrient supply, or in allocation to immunity. Instead, predictions in these other contexts require a mechanistic framework of competition between parasites involving host immunity and/or host resources.

### Section II: Within-host feedbacks driving divergent infection outcomes

To move toward creation of such a framework, we outline six models of within-host competition between parasites involving immunity, resources, and/or direct interference (Fig. 2). All potentially contain mechanisms for coinfection, single infection due to exclusion or priority effects (aka, alternative states or founder control), and no infection. Although details can vary, such divergent outcomes in these models follow general principles of species competition as outlined above. In essence, coinfection occurs when strong intraspecific competition (mediated directly or looped indirectly via ‘niche’ factors such as resources or immune cells) outweighs strong interspecific competition. Stronger intraspecific competition leads to net negative feedback on the feasible (’interior’) stable equilibrium, facilitating coinfection.

#### Two parasite – shared energy (2PE) or – shared immune cells (2PI)

Parasite species that share a resource (energy) or suffer attack from a shared immune system engage in exploitative or apparent competition, respectively (Fig. 2A,B). In exploitative competition (Fig. 2A), a parasite that can survive at the lower equilibrium energy (resource) level (*E\**) can outcompete the other. For instance, among two clones of a rodent malaria (*P. chabaudi*), the superior competitor for red blood cells (RBC) always excludes the inferior clone (De Roode *et al* 2005). Competition for a single, shared resource can lead to a competitive hierarchy among multiple parasite species. For instance, in a malarial-hookworm system, one Plasmodium species (*P. falciparum*) outcompetes hookworms which themselves outcompete another Plasmodium (*P. vivax*) for red blood cells (Budischak *et al* 2018). Analogous rules can characterize host systems with a shared immune system (Fig. 2B). In models, when two exponentially growing parasites share immune cells, the species that can withstand highest density of immune cells (*i.e.,* highest *I\**) wins via apparent competition (Fenton & Perkins 2010). In a possibly analogous experiment, immune-mediated interactions led to competitive suppression of an avirulent malarial clone, allowing the virulent clone to dominate (Råberg *et al* 2006). Therefore, exploitative competition for a single resource or apparent competition involving shared immune cells can lead to competitive exclusion alone.

Three aspects of within-host parasite biology can introduce coinfection or priority effects to pure resource or apparent competition. First, if some mechanism imposes self-limitation on both parasites, then they can coinfect even if they share only one immune niche. For instance, donor-controlled reproduction can create strong self-limitation on macroparasite density (Fenton & Perkins 2010). Second, non-linearity in immune clearance rates can generate self-limitation, particularly when clearance accelerates at higher parasite density (Fenton & Perkins 2010). Third, interference competition between parasites can facilitate priority effects or coinfection (Amarasekare 2002; Fig. 2A-B with mutually inhibitory effects between *P_1_* and *P_2_* [grey shading]). If the inferior energy competitor is superior at interference, priority effects ensue. Then, at high enough initial densities, the inferior energy competitor can win instead of being excluded.

However, if the interference also confers a benefit to the interacting species (e.g., if killing competitor larvae and consuming it increases per capita growth rate), then coinfection ensues (following Amarasekare 2002). These general principles for joint interference and exploitative competition (Fig. 2A) likely also apply to immune-mediated apparent competition, a possibility for future exploration (Fig. 2B).

#### Two parasite – two energy (2P2E; Resource ratio) or Two parasite – two immune cells (2P2I; Immune ratio)

When parasites (*P_j_*) simultaneously compete for two resource or energy sources (*E_i_*) via exploitative competition, infection outcomes can include coinfection and priority effects (Tilman 1982). Consider two parasite species competing for two substitutable resources (like in the resource ratio model: Fig. 2C). Here, coinfection minimally requires each species to trade off their requirements for each energy resource. Then sufficiently intermediate supply ratios must permit each single-species (‘boundary’) equilibria to fall within each competitors’ niches (enabling mutual invasibility). With both conditions met, coinfection arises if each parasite has larger impact on the resource to which its fitness is most sensitive (see Appendix Section 2). In testing the resource ratio model, Lacroix *et al*. observed a distinct competitive hierarchy, where one plant virus altered success of a competing cereal dwarf virus under nitrogen: phosphorus (N:P) supply. In co-inoculated plants, the cereal dwarf virus infection rates decreased with elevated *P* supply rate, while the addition of N significantly increased its’ infection rate (Lacroix *et al* 2014; Smith 2014). This suggests that, at the very least, N:P shapes how each parasite species differentially impacts their resources, setting the stage for coinfection.

Analogous rules likely apply to hosts with two types of immune response (2P2I, the immune ratio model; Fig. 2D). If parasites act as substitutable resources for two generalist immune responses, coinfection arises if each parasite also has larger impact on the immune response to which its’ fitness is most sensitive (Appendix Section 2; derived for case with parasite self-limitation). Modification of this 2P2I framework captures well-studied within-host interactions. First, the host can mount an independent, specialist immune response to each parasite (with no links between *P_i_* – *I_j_*, Fenton & Perkins 2010). Therefore, the separate dynamics of each immune-parasite pair determines coinfection. Similar results emerge for parasites separated spatially (e.g., in different host tissues: Cervi *et al* 2004; Karvonen *et al* 2006). Second, two parasites can each be attacked by specialist immune responses that inhibit each other. For instance, in mice, competing parasites interact *via* T-helper cells, where Th1 attacks intra-cellular malaria, Th2 attacks intestinal nematodes, but Th1 and Th2 inhibit each other (Griffiths *et al* 2015). Such interference creates negative feedback that can further enable coinfection (*e.g.*, links between *I_1_* and *I_2_* [grey shading]; Fenton & Perkins 2010).

#### Two parasite – immune cell – energy (2PIE)

Parasites can also engage in simultaneous exploitative and immune-mediated apparent competition (Cressler *et al* 2014; Ramesh & Hall 2023). Consider common variations of two competing parasites (*P_j_*) that steal energy (*E*) from hosts and immune cells (*I*) that kill them (2PIE; Fig. 2E-F; Table S1). In one, only parasites induce production of immune cells (2PIEi, induced immunity). In the other, energy is continuously allocated to maintain baseline immune function, even without parasites (2PIEc, constitutive immunity). This structure resembles a food web in which two prey species share a resource while attacked by a generalist predator (Holt *et al* 1994; Leibold 1996). This keystone predation (diamond) model anticipates the tradeoffs and niche dimensions that would govern exploitative and apparent competition parasites (Ramesh & Hall 2023). When combined, these forms of competition enable coinfection, priority effects, or competitive exclusion (see below Fig. 3; Ramesh & Hall 2023). Constitutive immunity (Fig. 2F) reduces opportunities (parameter space) for coinfection, lowering parasite burden while maintaining higher energy for other metabolic work (see below; Fig. 9).

The remainder of this review will focus on joint immune and energy (resource) competition in the 2PIE model. Most organisms have some type of (costly) immune defences to fight parasites that steal host energy (*reviewed in* Lochmiller & Deerenberg 2000; Zuk & Stoehr 2002). Furthermore, principles that govern strength of intra- *v* inter-specific competition in 2PIE models should apply to others. With the 2PIE model, then, we ask three questions: (a) What within-host feedback drives divergent outcomes of infection? (b) How does this framework offer new ways to resolve results in previous coinfection experiments (Fig. 1A-C)? (c) How can this framework guide future theory and experiments?

### Section III: A mechanistic framework linking feedbacks, trait ratios, and infection outcomes

In this section, we construct a framework for within-host competition of parasites that connects trade-offs, nullclines (niches), feedbacks, and traits. As we will learn, the 2PIE model follows the familiar tale of intra- vs inter-specific competition governing coexistence vs priority effects (given a feasible interior equilibrium; Fig. 1D). Although specifics between 2PIEi and the others (Fig. 2A-F) surely differ, this approach lays a general script to generate mechanistic *a priori* predictions and glean better understanding of coinfection dynamics from within-host niche models. Many of the mathematical details are found elsewhere (Appendix Section 1).

#### *The Model* (see also S1, Table S1)

*Growth rate of immune cells I* (Eq. 2A): In 2PIEi, immune cells (*I*) increase after attack on parasite *j* (*P_j_*) at rate 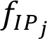, inducing consumption of 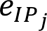 of energy, *E*, with conversion efficiency *e_I_* of energy into an immune cell after debiting loss rate *m*_I_. In 2PIEc, *I* also increases with allocation of *E* at baseline rate *a_b_*.

*Growth rate of parasite P_j_* (Eq. 2B): The two parasites consume host energy with feeding rate 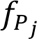 and energy per parasite conversion, 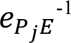. The parasites are lost due to attack by immune cells 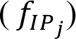 and die at (shared) background rate *m*_P_.

*Growth rate of energy*, *E* (Eq. 2C): The host consumes a resource *S* via a Monod function with maximal assimilation rate *f*_E_and half-saturation constant *h.* (This function merely pays homage to non-linear feeding behaviour of hosts following Cressler *et al* 2014). That resource, converted to energy within the host (*E*), is lost at fixed rate *r* for use by hosts (for metabolic needs). Its net production, then, is *f*(*S*) – *r E*. Additionally, host energy is consumed via induction from immune attack on parasites (proportional to the triple product *P_j_ E I*), and from consumption (theft) by parasites themselves. Hence, parasite *j* ‘consumes’ energy both indirect and directly described, a sum grouped below as *f_j_*(*I*). In 2PIEc, energy is also lost via allocation to baseline immune function at rate *a_b_*. The 2PIEi,c models are thus:

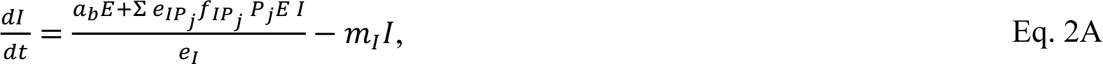

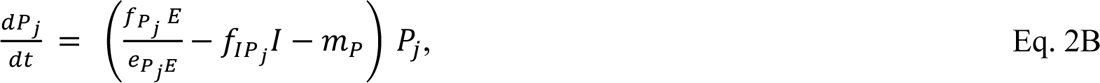

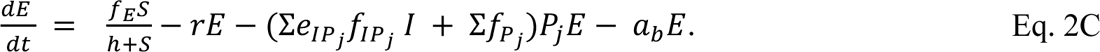

#### Outcomes of competition

Depending on their traits, competing parasites can successfully coinfect, singly infect (via competitive exclusion or priority effects), or produce no infection in a host. Such outcomes can be captured along gradients of, say, supply of resources to hosts, *S*, and feeding rate of parasites on energy within hosts, 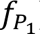 (Fig. 3A, Table S1). The lines within the 2D bifurcation diagram represent qualitative shifts of within-host composition determined by minimal requirements. We highlight two such shifts or ‘transcriticals’ (*T_1_* and *T_2_*) governing coinfection, but we build to it. Very low *S* prevents persistence of either parasite, resulting in no infection (a nutritional clearance; Fig. 3A, yellow). Increasing *S,* now supports only single infection by parasite 1 (*P_1_*) because it excludes the other via resource competition (*P*_2_, at *E-P_1_* [lighter] or *E-P_1_-I* [darker] ‘boundary equilibria’). Now, intermediate *S*, such that *T_2_* < *S* < *T_1_*, allows parasite 2 (*P_2_*) to invade that *P_1_* boundary and *P_1_* invasion into the *P_2_’s* boundary equilibrium (*E*-*P*_2_-*I*). Hence, that region represents mutual invasibility and coinfection (Fig. 3A, orange), situated between regions of competitive exclusion (‘single infection’). At higher *S*, *P_1_* is excluded allowing *P_2_* to win (*E*-*P*_2_-*I*; Fig. 3A, dark purple). Notice, increasing both 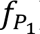 and *S* flips the geometry of those transcriticals. Now, higher *S*, such that *T_1_* < *S* < *T_2_*, does not allow *P_1_* invasion into the *P_2_’s* boundary equilibrium and vice-versa. Hence, this region represents mutual uninvasibility and priority effects aka founder control (Fig. 3A, grey; see Appendix S1 for details). The offsets show sample dynamics when hosts are exposed to concurrent infection, started at one or two initial densities, in each of these regions (Fig. 3B – F). In the no infection region, insufficient energy caused failed infection, even at high initial densities of parasites (Fig. 3D). In the single infection regions, the winner always excludes the other, even with initially low densities (Fig. 3A, E). In the coinfection region, all trajectories head toward a stable (interior) equilibrium (Fig. 3C). With priority effects, the parasite starting more densely wins and excludes the other (i.e., top: *P_1_* wins; bottom: *P_2_* wins; Fig. 3F).

#### Mechanism of competition: A within-host “assembly rules” and nullcline approach

These predictions hinge on intra- vs inter-specific competition. Intraspecific competition, mediated indirectly via host energy and immune cells, produces negative feedback loops while interspecific competition yields positive ones (Fig. 1D). When strength of intra-exceeds inter-specific competition, net negative feedback on the feasible interior equilibrium can enable coinfection (coexistence). Conversely, greater inter-than intra-specific competition generates net positive feedback that can produce priority effects. This section actualizes these concepts for 2PIEi using niches and nullclines.

##### Assembly rules

Broadly, coinfection requires a competition-resistance trade-off: the species that competes superiorly for energy (without immune cell) must remain more vulnerable to immune attack. It also requires intermediate resource supply, *S*, as consumed by hosts, *f*(*S*), falling in between two thresholds (described below: *T*_2_ < *f*(*S*) < *T_1_*). The assembly of a within-host ‘community’ of parasites can be understood via hosts exposed to (i) single species (Fig. 4A, 5A dotted and dashed lines) and (ii) both species (Fig. 4B, 5B solid lines). The single species ‘boundary’ equilibria provide insights into their minimum resource requirements of parasites for energy (*E_j_\**, akin to *R*\* *sensu* Tilman 1982) and of immune cells for the product of energy and parasites (*EP_j_\**). They also highlight maximal immune cell densities supported by each parasite (*I_j_\**, akin to *P*\* *sensu* Holt 1977). These quantities provide a start for assembly of coinfection.

**Figure 4:**
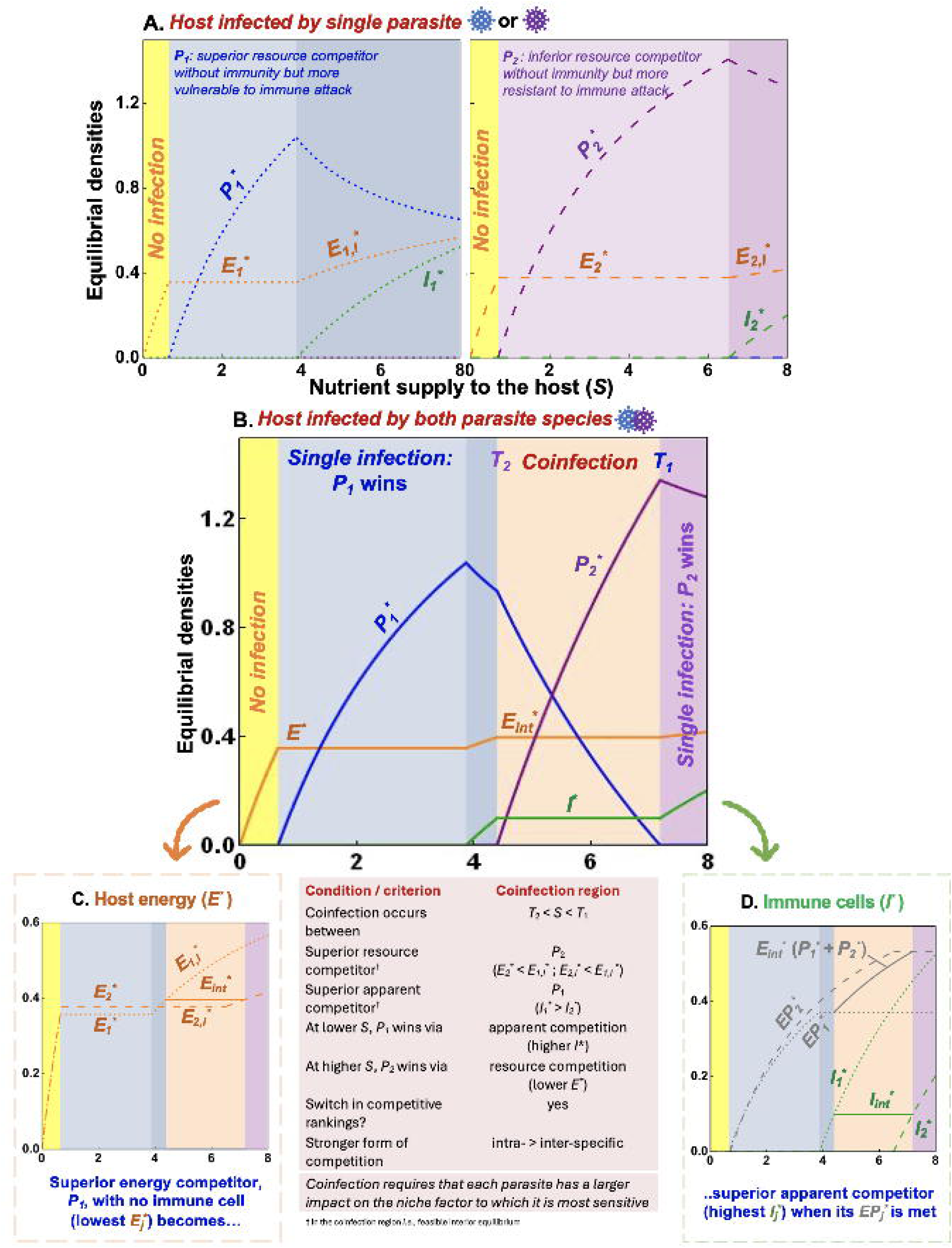
*Elements of coinfection (coexistence) in the 2PIEi model.* (A-B) Equilibrial densities along a gradient of nutrient supply (*S*) of (A) one single parasite, *P_1_* (blue region, tiny dash, left panel) or the other, *P*_2_ (purple, large dash, right) or (B) by both parasites (coinfection in orange) for a fixed feeding rate of 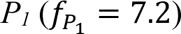. (C-D) A closer comparison of energy (*E*) and immune (*I*) quantities of each parasite provides insight into outcomes of joint resource (energy) and apparent competition. (C) *Energy competition*: Without immune attack, *P_1_* is the superior energy competitor (*E_1_\** < *E_2_\**) allowing *P_1_* to win (*sensu* the *R*\* rule). Mortality from immune activation weaken *P_1_*’s competitive ability, allowing *P_2_* to become the superior energy competitor (*E_1,I_*\* > *E_2,I_*\*). (D) *Immune-mediated apparent competition*: Immune cells proliferate when their minimum energy-parasite requirement is met (*EP_1_*\*: gray dotted; *EP_2_*\*: dashed). The winner of apparent competition (higher *I_j_*\*) supports more immune cells (here: less resistant *P_1_*). Red box: Conditions promoting coinfection.

**Figure 5:**
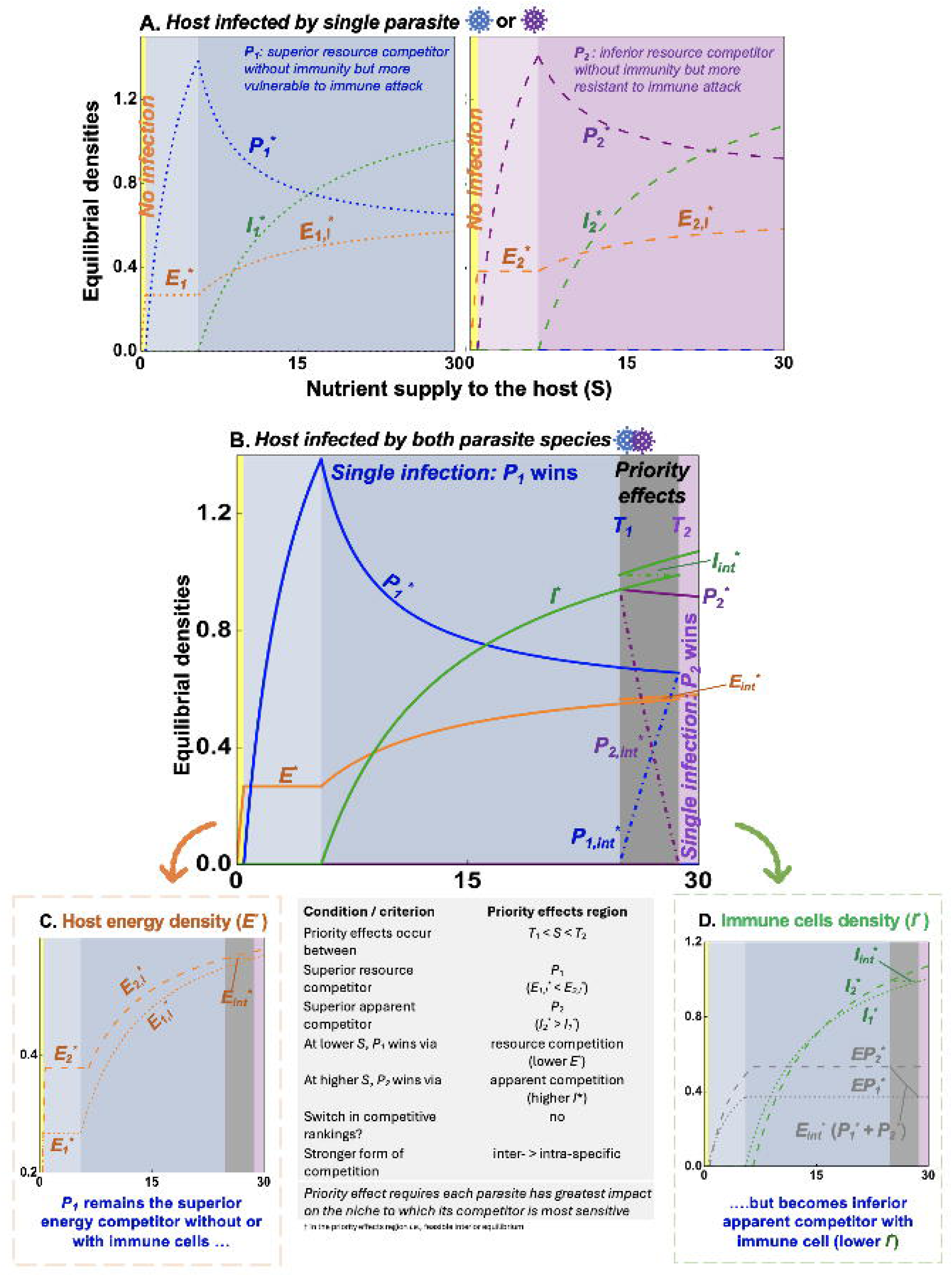
*Priority effects (founder control) in the 2PIEi model using assembly rules.* (A-B) Equilibrial densities along a gradient of nutrient supply (*S*) of (A) one parasite, *P_1_* (blue region, tiny dash, left panel) or the other *P*_2_ (purple, large dash, right) or (B) leading to priority effects (grey) for a fixed feeding rate of 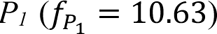. (C-D) Comparison of energy (*E*) and immune (*I*) quantities with each parasite singly provides insight into competitive outcomes. (C) *Energy competition*: Here, *P_1_* always remains the superior energy competitor with (*E_1_*\* < *E_2_*\*) or without (*E_1,I_*\* < *E_2,I_*) immune cells. (D) *Immune-mediated apparent competition*: Immune cells invade when the minimum energy-parasite requirement of the immune cells is met (*EP_1_*\*: gray dotted; *EP_2_*\*: dashed). In the region of priority effects, more resistant *P*_2_ wins via apparent competition (since *I_2_\** > *I_1_*\*). Grey box: Conditions promoting priority effects.

First, the route to single infection follows resource- and immune-based assembly rules along a nutrient supply gradient, *S* (Figs. 4B, 5B). Very low nutrient supply (*S*) meets neither parasite’s minimum energy requirements (*E*\* < *E_j_\**). Hence, very low *S* prevents infection (i.e., clearance via starvation; yellow). However, with high enough *S*, *E\** increases, eventually meeting that minimum (*E_j_\**). Then, *P_j_* can invade and increase with *S* (pinning *E* at *E_j_\**; *E-P_j_* regions in lighter). Eventually, *P_j_*’s density crosses a minimal threshold (*EP_j_*\*) that induces immune activation (darker colors). With higher *S* still, immune attack reduces density of *P_j_* freeing up more energy (where energy with immune cells is denoted by *E_j,I_\**). While both parasite species follow similar assembly rules, they differ in a key trade-off: *P_1_* is the superior competitor without immune cells (*E_1_\** < *E_2_\**) but is more vulnerable to immune attack 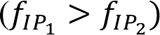. That competition-(resistance trade-off provides a first prerequisite for coinfection.

Comparison of those key energy and immune quantities yields more conditions for coinfection (Fig. 4B). Along the nutrient gradient, *S*, the more vulnerable parasite, *P_1_* invades (*E-P_1_*). It outcompetes (excludes) the more resistant parasite, *P_2_*, via resource competition (since *E_1_\** < *E_2_\**). At some *S* following immune activation (*EP_1_\**), *P_1_* becomes the superior apparent competitor (*I_1_\** > *I_2_\**) but inferior energy competitor (*E_2,I_\** < *E_1,I_\**) – a shift in competitive dominance. Then, with further *S* still, the *E-I* niche environment set by *P_1_* enables *P_2_* to invade (at *T_2_*: orange).

In a window of coinfection, the within-host niche environment remains constant while parasite densities shift (Fig. 4B). Here, energy and immune cells stay at *E_int_*\* and *I_int_*\*, respectively. However, with higher *S*, the structure of this community shifts, with *P_2_* increasing while *P_1_* declines. With this shift, the energy and parasite density needed to keep the immune system activated, *E_int_*\*(*P_1_*\*+ *P_2_*\*), increases until *P_2_* could support immune activation alone (at *EP_2_\**; Fig. 4D). Broadly, in this region, the more vulnerable *P_1_* remains the superior apparent competitor (Fig. 4D, *I_1_* > I_2_*\*) but weaker competitor for energy (Fig. 4C, *E_2_*\* < *E_1,I_*\* shifts to *E_2,I_*\* < *E_1,I_* *). Conversely, the more resistant *P_2_* is the superior energy competitor but weaker apparent competitor. This biology means that the more vulnerable parasite species also produces more immune cells, the niche factor to which it is most sensitive (see below). That combination imposes a brake on its own growth rate (negative feedback), thus promoting coinfection. With *S* past this threshold (*T_1_*), *P_2_* excludes *P_1_* (via resource competition: *E_2,I_\** < *E_1,I_\**) but more immune cells lower *P_2_*’s density (purple, Figs. 5B-E).

Priority effects can arise with different trait combinations for the two parasites (here: at higher 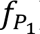; Fig. 5B). Along a gradient of nutrient supply to hosts, *S*, the region of priority effects is approached similarly to the coinfection case: first the more vulnerable parasite invades (when its *E_1_\** is met), then the immune system is activated (when its *EP_1_\** is reached). However, *P_1_* always remains the superior energy competitor, unlike the flip seen before the coinfection window for coinfection (Fig. 5C). Past *T_1_*, more resistant *P_2_* cannot invade until *T_2_* is crossed; with higher *S*, more resistant *P*_2_ excludes *P_1_* via apparent competition (since *I_2_*\* > *I_1_*\*; Fig. 5D). Biologically, this means that the more resistant species produces more immune cells, the niche factor to which their competitors are most sensitive (see below), creating positive feedback and thus priority effects. With decreasing *S*, *P_1_* could not invade until *T_1_* was crossed. Hence, the interior equilibrium in the priority effects window is unstable (a saddle), separating two stable states, dominance by *P_1_* (at its *E-P_1_-I* boundary equilibrium) or by *P_2_* (at *E-P_2_-I*).

##### A nullcline (niche) interpretation

Nullclines help to characterize the niche environment that enables it coinfection. At a given nutrient supply *S*, nullclines denote conditions in which a parasite (*P_j_*) or niche component (energy, immune cells) neither grows nor declines. Combinations of energy (*E*) and immune (*I*) cells that fall to the right of a parasite’s nullcline (higher energy, lower immune cells) sit within its fundamental niche (Fig. 6A*; P_1_*, blue shading; *P_2_*, purple). Then, the impact of each parasite on their within-host niche determines *I* and *E* nullclines. The nullcline for *E* is the combination of *P_1_* and *P_2_* that ‘consumes’ all production of energy at the interior (*E_int_*\*). Below this line, energy grows (d*E*/d*t* > 0, yellow in Fig. 6B). The *P*_j_-axis intercepts 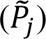 note the density of each parasite, when alone, that consumes all net energy production. The nullcline for *I* is the combination of *P_1_* and *P_2_* that meets the minimal *EP*\* needed to maintain its activation. All points above the line increase production of immune cells (d*I*/d*t* > 0; green in Fig. 6B). At each intercept 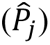, the parasite alone supports immune activation (at *EP_j_*\*) given *E_int_*\*.

**Figure 6:**
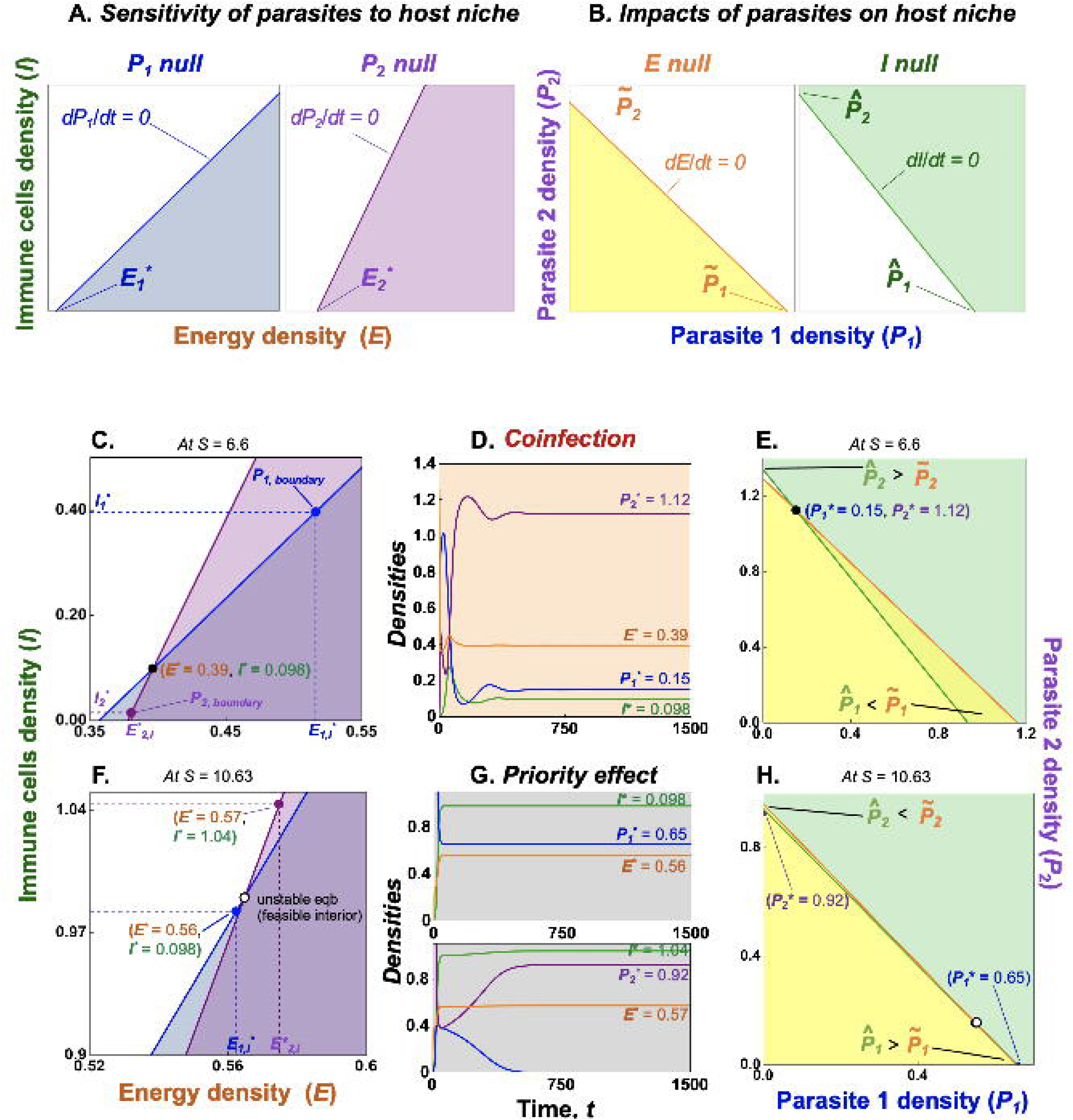
*Interpretation of coinfection (coexistence) and priority effects (alternate stable states) in the 2PIEi model using nullclines.* (A) *Sensitivity to their niche*: Densities of energy (*E*) and immune cells (*I*) at which parasites 1 (*P_1_*) and 2 (*P_2_*) show zero growth (*dP_j_*/dt = 0). Combinations below these nullclines (higher energy, less immune attack) fall in the fundamental niche of each parasite. (B) *Impacts on their niche:* densities of parasite 1 (*P_1_*) and 2 (*P_2_*) below the energy nullcline (*dE*/dt = 0, yellow) or above the immune (*dI*/dt = 0, green) nullcline lead to increases in both. At a given *S*, the outcomes of within-host competition can also be visualized at the intersection of (C,F) parasite or (E,H) energy-immune cell nullclines involving the (D, G) equilibrium denoting coinfection (black, closed circle: C,D,E) or priority effects (white, open circle: F,G,H). (See text and Appendix S1, Tables S1,2 for more details).

*Coinfection* (Fig. 6C - E; black, closed circle): Co-infection requires that parasite nullclines cross, niche nullclines cross, and nutrient supply is intermediate. The nullclines of the parasites cross in *E-I* space because of their competition-resistance trade-off (Appendix section S1). The shallower slope of less resistant *P_1_*’s nullcline means it is more sensitive to *I* while more resistant *P_2_* is more sensitive to *E*. When they coinfect, both parasites set their relevant single-species (boundary) equilibria within the fundamental niche of the other. Hence, each single-parasite environment can be invaded by the other (Fig. 6C; *P_1_* equilibrium [blue dot] falls within *P_2_*’s niche [purple shaded] and vice-versa). Now *P_1_* is the superior apparent competitor (*I_1_* > I_2_\**) while *P_2_* is the superior energy competitor (i.e., has lower *E*\*: *E_1,I_ * > E_2,I_* * [as shown] or *E*_2_*; Fig. 4C). Meanwhile, the *I* nullcline is steeper in *P_1_-P_2_* space (Fig. 6E). Hence *P*_1_ has larger impact on immune cells while *P_2_* has larger impact on energy. Therefore, each parasite exerts largest impact on the niche factor to which it is most sensitive. Based on the nullcline intercepts, coinfection requires that the interior niche would support surplus 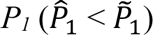 but deficient 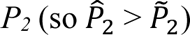 than needed to meet immune system’s *EP\** requirement (Fig. 6E; when 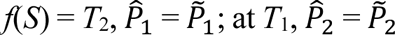). The coinfection equilibrium, then, combines densities of the superior apparent (*P_1_*) and energy competitor (*P_2_*) that consumes all net energy production while meeting minimal requirements of immune cells – eliminating those surpluses and deficits.

*Priority effects* (Fig. 6F-H; white, open circle): During priority effects, those three elements change. First, while parasite nullclines still cross (due to the competition-resistance trade-off), now both parasites (*P_j_*) alone create uninvasible immune (*I*) *–* energy (*E*) niche environments (Fig. 6F; *P_1_* blue dot outside *P_2_* purple shaded niche and vice-versa). Furthermore, in this region *P_1_* is always the superior energy competitor, without (*E_1_ * < E_2_* *) and with immune cells (*E_1,I_ * < E_2,I_*\*), while *P_2_* becomes the superior apparent competitor (*I_2_* > I_1_\**; Fig. 6F). Second, the *E* nullcline is now steeper than the *I* nullcline (Fig. 6J). Hence, impacts flip: less resistant *P_1_* has larger impact on *E* while more resistant *P_2_* has larger impact on *I*. Therefore, each parasite has a larger impact on the niche factor to which its competitor is most sensitive. Thus, the system would provide excess of the more resistant parasite 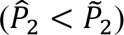 and a deficiency of the vulnerable one 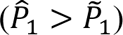 enabling priority effects.

#### Factors enabling coinfection: Feedback loops and trade-offs of traits

Outcomes of coinfection v priority effects depend on feedback and traits that govern them. At the most fundamental scale, co-infection must be ‘feasible’ (set by an environment supporting both species) at an equilibrium having net negative feedback. Such net negative feedback emerges when intraspecific competition exceeds interspecific competition. That competition depends on loops connecting parasites to their niche (Fig. 7A, S1). Loops are chains of interactions linking increased density of a species to growth rates of others, eventually returning to that species (Puccia & Levins 1991). For instance, interspecific competition can be traced starting with an increase in *P_1_*. (i) An increase in *P_1_*’s density can ‘*fuel immune cells’* that suppress its competitor, freeing up energy resources, thereby increasing its own growth rate via higher ‘births’. (ii) *P_1_* can also ‘*starve immune cells’* by reducing *P_2_*’s density via consumption of *E*. Through both chains of interactions, *P_1_* indirectly benefits from an increase in its own density via gains in energy, hence birth rate, or lower death rate, respectively. Those two positive (destabilizing) loops then push against two negative loops, (iii and iv) “*P_i_ is attacked, P_j_ eats*”. These latter loops add the stabilizing, consumer-resource-like, intraspecific competitive interactions within which each parasite is enmeshed. For instance, (iii) a small increase in *P_1_* increases immune attack, thereby reducing *P_1_* (*P*_1_-*I* loop) while *P_2_* is slowed by resource consumption (*P_2_*-*E* loop). Then, (iv) those roles reverse, *i.e.*, resources brake *P_1_* while the immune system slows parasite *P_2_*. Summed, those two loops (iii and iv) jointly determine the amount of intraspecific competition (negative feedback). Then, the 2PIE model follows the recognizable script of intra- vs inter-specific competition governing coexistence vs priority effects. If the strength of intraspecific competition loops (iii + iv: negative) exceeds that of interspecific loops (i + ii: positive), net negative feedback leads to coinfection. If, instead, interspecific exceed intraspecific loop strength, net positive feedback generates priority effects.

**Figure 7:**
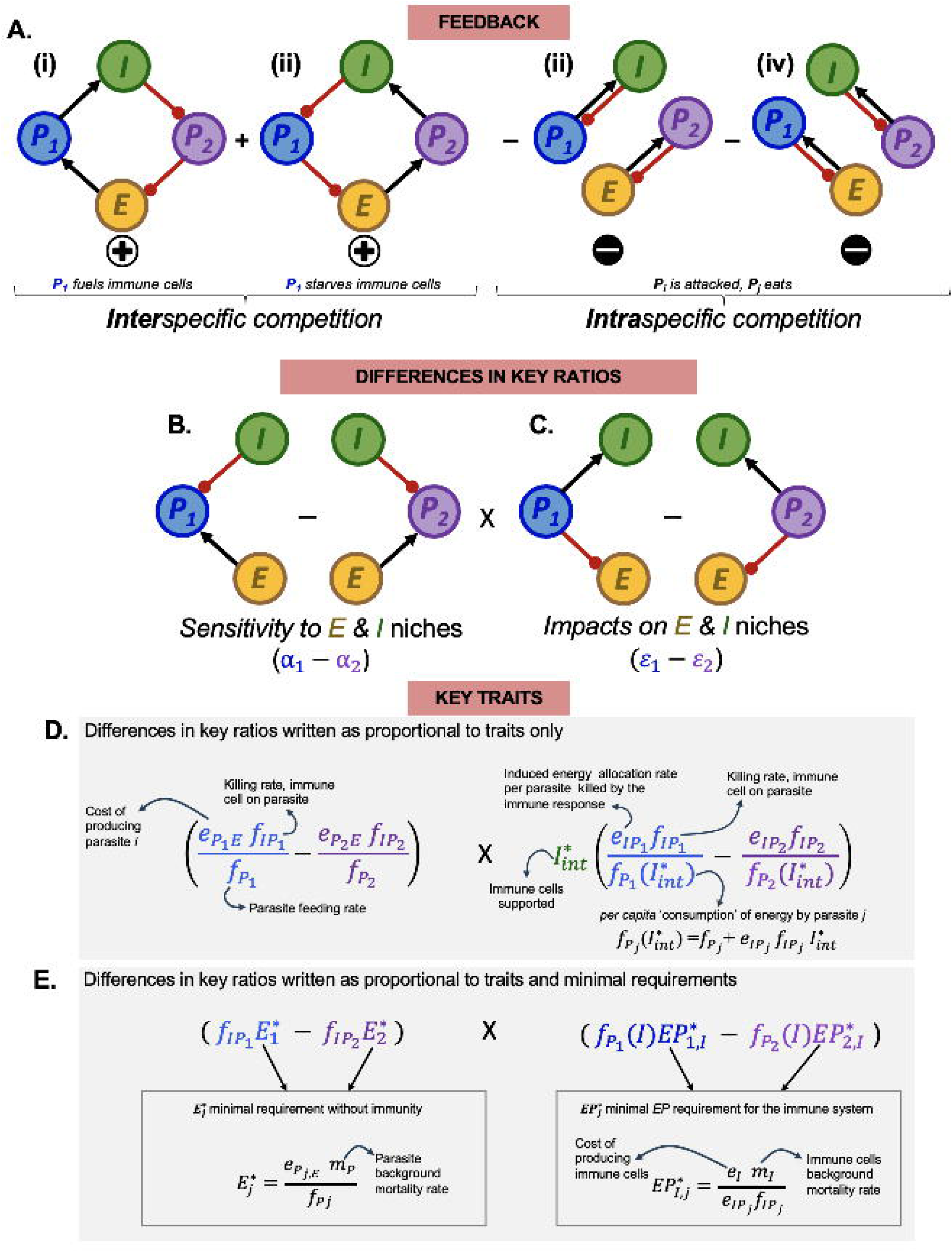
*A mechanistic framework for within-host parasite competition, linking feedback, trait ratios and key traits in the 2PIEi model.* (A) *Feedback:* Coinfection v priority effects is governed by the sum of level four feedback loops (*F_4_*); from *L-R*, two destabilizing, positive (+), interspecific competition loops and two stabilizing, negative (-), intraspecific competition ones. *P_i_* benefits from an increase in its density (positive feedback, interspecific) as it (i) directly “*fuels*” or (ii) indirectly “*starves*” immune cells but is restrained (negative feedback, intraspecific) by (iii) and (iv) “*P_i_ is attacked, P_j_ eats*” loops (the product of binary *I-P_i_* and *P_j_-E* consumer-resource-like loops). (B) *Differences in ratios of key traits:* With some rearrangement, these feedback loops correspond to ratios of key quantities made of key traits. These ratios encapsulate differences in how competing parasites are (B) *sensitive to* (*α*_1_ – *α*_2_) and have (C) *impacts on* (*ε*_1_ – *ε*_2_) immune cells and energy. Those differences in ratios can be written proportional to and measured either as a combination of (D) traits or as (E) traits and minimal requirements. For instance, (E) *α_j_* is proportional to the product of killing rate of immune cell on parasite *j* 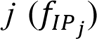 and the minimal energy requirement of parasite *j* (*E_j_*\*), and *ε_j_* is proportional to the per capita ‘consumption’ of energy by parasite *j*, 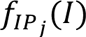, times minimal energy and parasite needed for immune activation (*EP_j_\**).

With some rearrangement, these intra- and inter-specific loops governing feedback can more intimately connect to the niche environment and assembly rules revealed above (Fig. 7B, S1). The first involves the difference in ratios of sensitivity of each parasite to immune cells, *I*, vs. energy, *E* (denoted by *α_j_*; following Pásztor et al 2016). The second involves the ratio of impacts of each parasite on *I* vs. *E* (*ε_j_*). Both ratios can be written as proportional to ratios of traits or traits and minimal requirements (Fig. 7D and E, respectively; see details in Appendix Section S1). Coinfection requires that each parasite has a larger impact on the niche factor to which it is most sensitive. If the more vulnerable *P_1_* is most sensitive to and has highest impact on *I*, then *α_1_* > *α_2_* and *ε_1_* < *ε_2_*. Such symmetry in ratios generates net negative feedback. For priority effects, the competition-resistance trade-off still yields *α_1_* > *α_2_*. However, because *P_1_* has highest impact on *E* (to which *P_2_* is most sensitive), the impact ratios flip, *ε_1_* < *ε_2_*. This asymmetry in sensitivity vs. impact ratios produces net positive feedback.

### Section IV: A focus on the traits and quantities to measure in the future

Presently, most within-host competition experiments observe patterns of coinfection, then infer mechanisms. The next phase of experimentation should shift toward feedback, minimal requirements, and traits of mechanistic niche models (like 2PIEi). Above, we learned that coinfection requires that each parasite has greater impact on the niche factor to which it is most sensitive (generating net negative feedback); with priority effects, each parasite has greatest impact on the niche factor to which its competitor is most sensitive (yielding net positive feedback). Future research on coinfection dynamics could aim to quantify the traits and minima’s underlying these sensitivity and impact frameworks.

1. *Quantifying traits:* The (a)symmetry of the *sensitivity-impacts* ratios governing coinfection and priority effects can be better understood by delineating the combination of traits (Fig. 7D, Appendix Section S1). For instance, the *sensitivity* ratios are governed by killing rate of immune cells on parasites 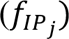, feeding rate of parasites 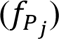, and the cost of producing a new parasite 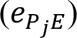. On the other hand, the *impact* ratios depend on how attacked parasites induce energy allocation to immune cells 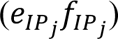 and the per capita ‘consumption’ of energy by parasite *j* directly and indirectly 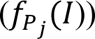.
2. *Quantifying key traits and minimal requirements:* Experimental tests of *sensitivity-impact* ratios could also centre on measurement of key quantities such as the minimal requirement of each parasite (*E\**) for energy and of the immune system for activation (*EP*\*; Fig. 7E, Table 1C). For instance, the difference in sensitivity ratios boils down to killing rate of immune cells on parasites, 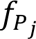, and each parasite’s minimal energy requirement without immune cells, *E_j_*\* Similarly, the difference in impact ratios depend on per capita ‘consumption’ of energy by parasites, 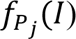, and the immune cells’ minimal energy-parasite requirement *EP_j_\** (Appendix S1). These traits and quantities could be estimated via experiments (in principle).

Despite this call for a focus on traits and minimal requirements, we acknowledge inherent challenges in measuring them. To move forward, experimentalists can leverage interdisciplinary methodologies. For instance, measurement of state variables like parasite (*P_j_\**), energy (*E_j_\**), or immune densities (*I_j_\**) over time can allow estimation of parameters of the model. Alternatively, traits could be measured with radioisotope labelling of resources to track parasite feeding rate (Gomez-Amaro *et al* 2015), live imaging to track immune killing rate (Galli *et al* 2021), ICP-MS to measure energetic content of infected host (Cassat *et al* 2018), etc. As a pay-off, the combination of trait measurements and models generates *a priori* predictions of infection outcomes and the feedback mechanisms that govern them (notably demonstrated in Budischak *et al* 2015; Griffiths *et al* 2015). Ultimately, such approaches may catalyse a new wave of theory-grounded, niche-mechanistic combinations of modelling and experimentation.

### Section V: Explanations for divergent infection outcomes using 2PIE

The model of exploitative and apparent competition between parasites, 2PIEi,c, makes predictions (Fig. 8A, 9) that can help to contextualize and interpret previous experiments (Fig. 8B-F). In this next section, we envision how 2PIEi could produce these various outcomes, backing-out mechanism *post hoc*. Thus, 2PIEi offers a way to potentially resolve otherwise seemingly inconsistent experimental outcomes. Then, it can guide creation of new coinfection models tested in future experiments. Ideally, such efforts would start with parameterized predictions that test divergence of infection outcomes *a priori*.

**Figure 8:**
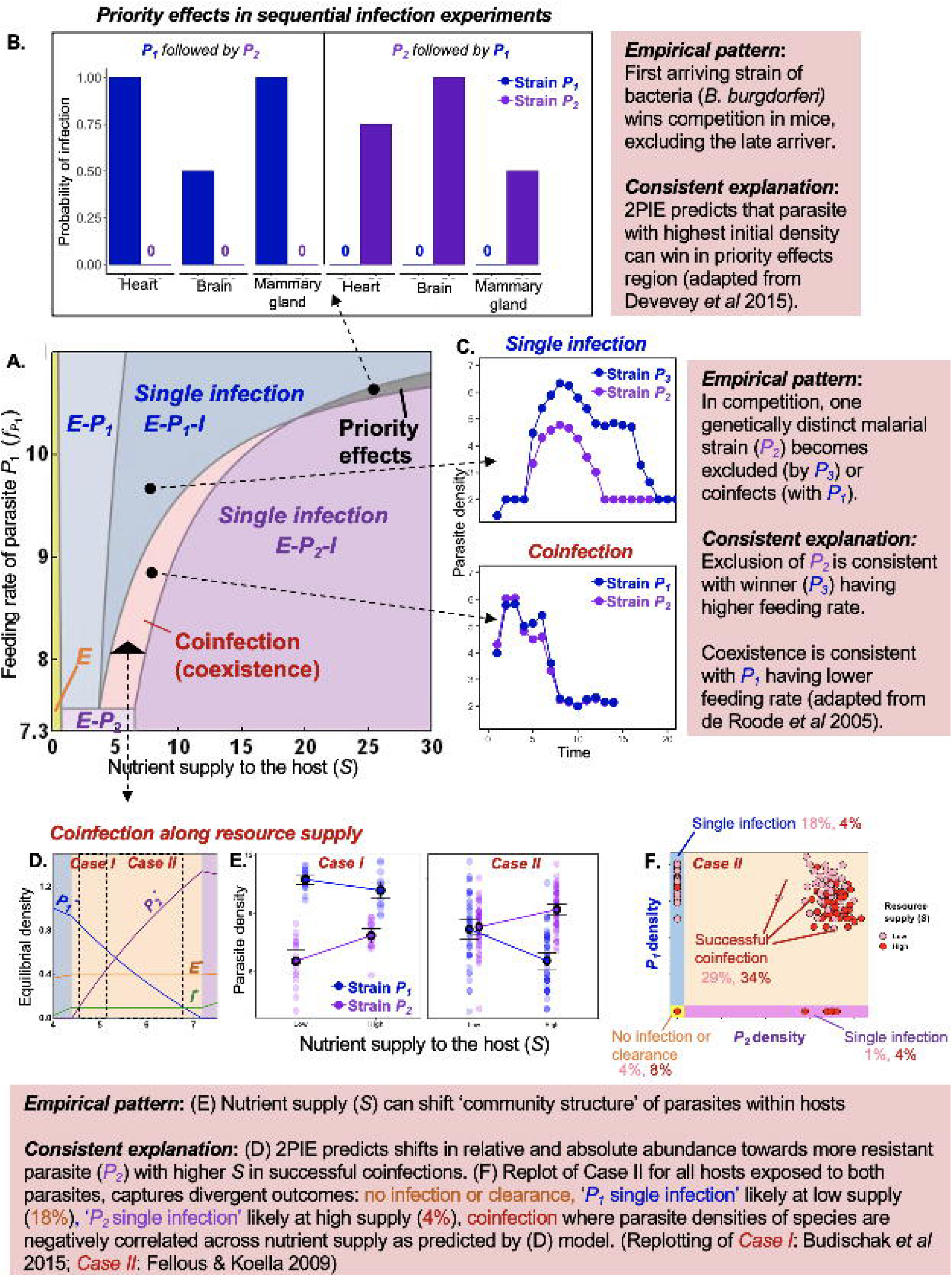
*Coinfected or not?* Assuming a competition-resistance trade-off between competitors, (A) the 2D bifurcation plot of resource supply to host resources (*S*) and feeding rate of parasite *P_1_* on host energy 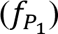 captures divergent infection outcomes in 2PIEi (as in Fig. 3): single infection either via priority effects (grey), or competitive exclusion (blue: *P_1_* wins; purple: *P_2_* wins), coinfection (orange), or no infection (yellow). (B) *Priority effects* in mice parasitized by strains of a bacterium (*B. burgdorferi*), where the first invader excludes the later. (C) *Competitive exclusion:* A malarial strain (*P_2_*; purple) is excluded by one (*P_3_*) or *coinfects* (coexists) with another strain (*P_1_*; blue). (D-F) *Coinfection community structure:* Coinfecting parasites can shift relative and absolute abundance within hosts with increasing resource supply (*S*). (D) In 2PIEi, increasing nutrient supply favours more resistant *P_2_* over the superior *P_1_*. (E) Empirically, shifts in community structure arise in mice coinfected by two species of gastrointestinal worms (case I) and a mosquito larvae infected by a microsporidian (case II). (F) Re-examination of all outcomes of competition from Case II suggests that variation in infection outcomes could arise when individuals assimilate different amounts of food. Small deviations among individuals could drive large variation in infection outcomes.

#### Coinfected or not? What within-host factors facilitate or inhibit coinfection? *(Fig. 8B, C)*

##### Priority effects

*Theory*: Stronger inter-than intra-specific competition ensures priority effects (via net positive feedback) where the parasite with sufficiently high initial dose wins (Fig. 8A, B; Table 1B).

*Empirical evidence:* In a rare, unequivocal demonstration of priority effects, the order of arrival of bacterial (*B. burgdorferi*) strains determined which won and excluded the other within a host mouse (Fig. 8B). Immune response likely did not explain the priority effects (Devevey *et al* 2015), so future work can pinpoint mechanisms (e.g., resource competition, interference, etc.) facilitating them (Fig. 2B; Table 1B).

*Guiding future experiments*: Varying initial parasite dose (dose-dependent assays) provide on possible way to delineate coinfection vs priority effects. When parasites coinfect, dose does not change outcome, despite varying initial *P_2_* and constant initial *P_1_*. With priority effects, sufficiently high initial density determines winner (Table 1B). Such an experiment would reveal whether competitive exclusion, or priority effects led to single infection. Even better, measurement of key quantities involved in *sensitivity-impact* ratios can delineate mechanism (e.g., superior resource *v* resistant competitor as in Fig. 5).

##### Competitive exclusion

*Theory:* Differences in parasite traits can separate exclusion from coinfection. For example, for a given nutrient supply to hosts, a parasite with higher feeding rate (all else equal), can exclude its competitor resulting in single infection (Fig. 8A, C). In contrast, one with lower feeding rate may successfully coinfect. *Empirical evidence:* Mice infected with malarial strain *P_1_* or *P_3_* (blue) were allowed to compete pairwise against a common malarial strain *P_2_* (purple; Fig. 8C; De Roode *et al* 2005). In the *P_3_* - *P_2_* pairing, regardless of order or arrival or delay between infections, *P_3_* competitively excluded *P_2_* (Fig. 8C).

*Guiding future experiments:* Higher competitive ability of *P_3_* may reflect higher feeding rate on host energy. In contrast, if strain *P_1_* had lower feeding rate (hence less competitive), it could stably coinfect with *P_2_* (Fig. 8C). Such hypotheses about resource acquisition traits could be tested in the future (Table 1B).

#### Coinfection hierarchy and community structure *(Fig. 8D-F)*

*Theory*: When parasites coinfect, nutrient supply to hosts (*S*) can mediate densities of the within-host energy and immunity, and hence relative and absolute densities of parasites (Fig. 8D). Thus, nutrient supply can shift this ‘community structure’ of parasites within hosts. For instance, increasing nutrient supply favours the more resistant *P*_2_ over the less resistant *P_1_*.

*Empirical evidence:* This prediction could explain shifts in community structure of parasites in two different systems (Fig. 8E). The first arose with mice infected by two species of gastrointestinal worms (case I; Budischak *et al* 2015); the second used mosquito larvae infected by a microsporidian and protozoan parasite (case II; Fellous & Koella 2009). Both demonstrate how changing nutrient supply (*S*) favours one species (purple; positive slope) over the other (blue; negative slope; Fig. 8D). In both cases, relative abundance of coinfecting species can shift (or not) depending on *S*, too. However, future trait measurements would need to establish that higher *S* favoured the more resistant parasite as 2PIE predicts (Table 1C).

*Guiding future experiments:* Much may be learned about coinfection and nutrient supply with focus on individual hosts (*reviewed in* Ezenwa 2021). Hosts can exhibit high intra- and inter-individual variation in infection outcomes (Merill & Cáceres 2018). To illustrate, parasite densities in the mosquito case ranged among all individuals from coinfection to single infection via exclusion / priority effects to no infection (see reference to shaded regions in Fig. 8D *v* 8F; replotting of Fellous & Koella 2009). Such variation among hosts could arise from individual differences in resource acquisition. If so, individuals nominally fed the same amount (in a treatment) may fall functionally along different supply points (like in Fig. 8D). Individuals consuming less resources would favour the less resistant parasite (*P_1_*) while those eating much more would favour the more resistant one (*P_2_*) - yielding exclusion in both cases (Fig. 8D, F). The remaining hosts became coinfected, with parasites reaching different densities among hosts. Future, experimental tests can test for the full range of possible infection outcomes (and hence ‘assembly rules’ leading to coinfection outcomes) at the individual scale, say, along broader nutrient gradients (Table 1C).

Furthermore, niche-based insights can guide more predictive experiments at the within-host scale which can then be scaled to the population linking within-to between-host dynamics. For instance, fluctuation in nutrient supply to host could shift competitive outcomes within hosts that then alters multi-parasite outbreaks at the population scale (Hite & Cressler 2018; Ezenwa & Jolles 2011) or ecosystem processes (Kendig *et al* 2020).

#### Coinfection burden (Fig. 9)

Finally, some hosts present higher coinfection burden (total density) than others, hinting at intrinsic host resistance via immune clearance. How does variation in immune response govern coinfection burden?

*Theory*: In comparison to the model with only immune induction (2PIEi), baseline energy allocation to immunity squeezes parameter space for coinfection and reduces parasite burden (2PIEc, constitutive immunity; Fig. 9A-B). Despite the higher allocation (via *a_b_*), reduced burden allows the infected host to maintain more energy for other metabolic work (Fig. 9A-B). At lower nutrient supply (*S*), ‘no infection’ by parasite shifts from nutritional clearance (*E*, yellow; Fig, 9A) to combination of immune and nutritional form of clearance (*E-I*, green; Fig, 9B). The 2PIEc model also predicts that increasing baseline allocation favours the more resistant *P_2_* over *P_1_*, eventually excluding it (Fig. 9E).

**Figure 9:**
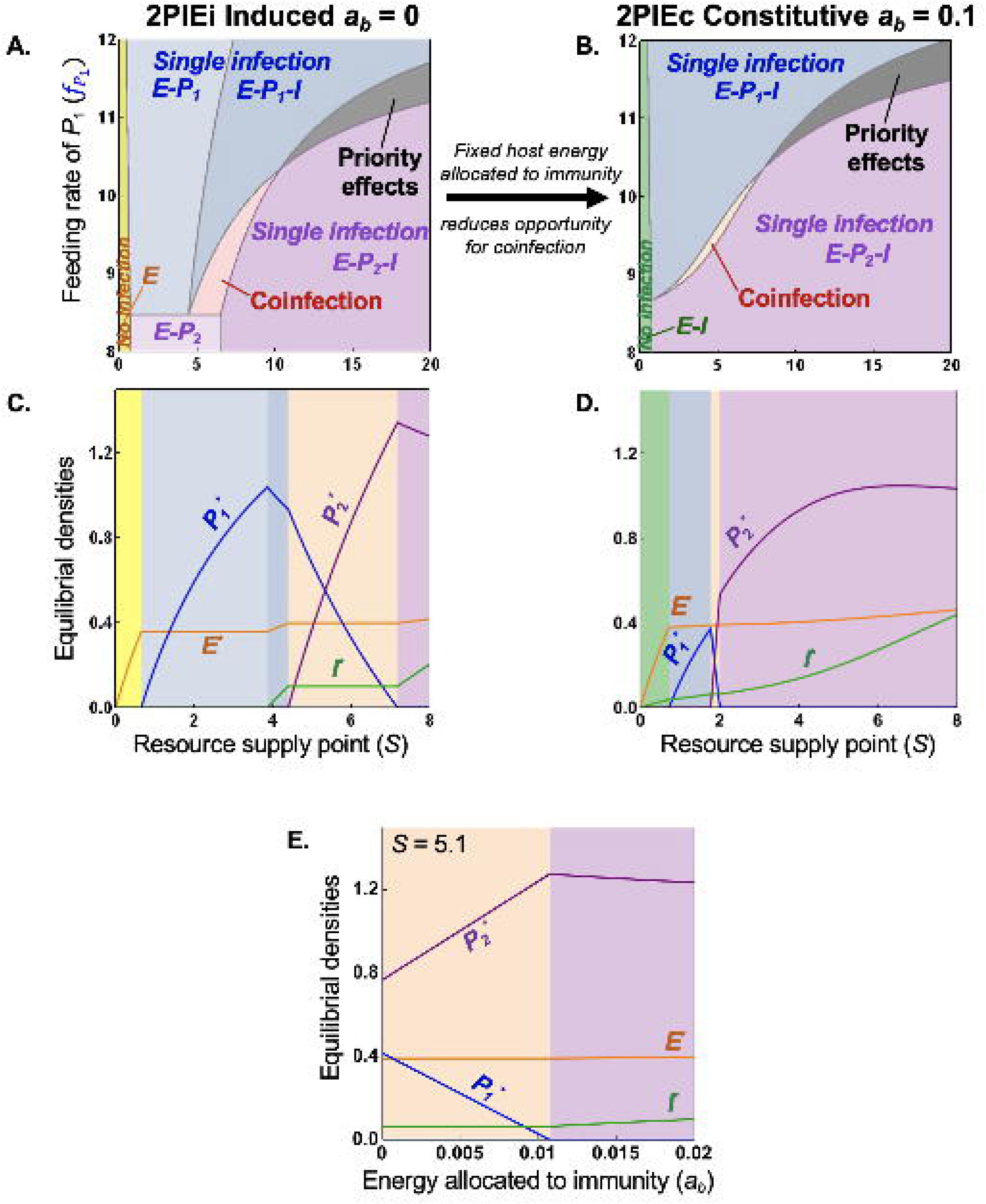
*Coinfection burden:* The burden of parasites that infect a host can depend on baseline energy allocation to immunity (*a_b_*). (A) vs. (B): Higher *a_b_* squeezes parameter space for coinfection (2PIEi, induced *v* 2PIEc, constitutive immunity, respectively). (C-D) It also reduces density of competing parasites. Reduced burden allows the host to maintain slightly more energy for other metabolic work, potentially improving host health. (E) Increasing *a_b_* favours the superior energy competitor (more resistant *P_2_*) over the superior apparent competitor (*P_1_*), eventually excluding it. By implication, differences in *a_b_* can lead to varying (co)infection burden among hosts.

*Empirical evidence*: Consistent with these predictions, plant hosts treated with immune-signalling hormone experienced a lower prevalence of a less aggressive parasite, increased burden of infection by a more aggressive parasite, and experienced fewer co-infections (Halliday *et al* 2018). Thus, hosts with higher allocation to immunity can resist infection more than those with lower allocation, leading to variation among hosts in infection burden (all else equal).

*Guiding future experiments:* First, one could test such predictions using strains with immune suppression of specific genes or metabolites involved in constitutive immunity (like in Chen *et al* 2005). That could shift competition from a 2PIEc framework (with *a_b_* > 0) to a more 2PIEi like one (with *a_b_* = 0 at the extreme). Another test could involve measuring competitive outcomes in host genotypes differing allocation to baseline immunity (Fuess *et al* 2021; Table 1D).

### Section VI: Conclusion

Why do divergent infection dynamics arise within a host? Competing parasites within a host can coinfect (coexist), singly infect (through exclusion, or priority effects), or become cleared (no infection; Fig. 1, 3). Those differing outcomes significantly affect health of individual hosts and even alter population-level disease outbreaks (Mideo *et al* 2008). Such divergent infection outcomes ought to stem from niche interactions, *i.e.,* with the resources that parasites steal from hosts and the immune cells that kill them (Cressler *et al* 2014, Graham 2008). Yet, despite two decades of empirical focus on infection dynamics, many of these studies present a collection of disparate results without a synthetic glue. Clearly then, we need a comprehensive framework that explains why parasites coinfect and why they might not.

Here we build a within-host framework of parasite competition based on ecological theory (Fig. 2). Using a two parasite – immune cells – energy model (2PIE), we illustrate general principles governing parasite competition via join exploitative and immune-mediated apparent competition (Ramesh & Hall 2023). Those forms of competition underlie a within-host framework that connects infection outcomes to competitive abilities, nullclines (niches), feedbacks, and traits (Figs. 3 – 7; see details in Appendix Section S1). Notably, we delineate how the interplay of three quantities – minimum resource requirements of (1) parasites for energy (*E_j_\**, akin to *R*\*), and of (2) the product of energy and parasites (*EP_j_\**) for immune cells, and (3) maximal immune cells supported by each parasite (*I_j_\**) – provide a start for assembly of coinfection.

We show that coinfection requires that each parasite exerts greater impact on its more sensitive niche factor; that arrangement introduces net stabilizing, negative feedback. With priority effects, each parasite more strongly impacts the factor to which its competitor is most sensitive, leading to net destablizing, positive feedback (Fig. 3, 4).

That framework also provides mechanistic insights into and explanations for some experimental results (Fig. 8-9). First, it explains two ways to hinder coinfection: priority effects can favour early invaders, while competitive exclusion always inhibits one parasite (arising via *e.g*., fast feeders at low nutrient supply; Fig. 5, 8B-C). Second, increasing nutrient supply in the coinfection region can favour the more resistant over the less resistant parasite, shifting community structure along nutrient gradients (Fig. 8D). Finally, greater investment in constitutive immunity squeezes opportunity (parameter space) for coinfection and reduces parasite burden, freeing up energy for metabolic needs (Fig. 9). Together, the 2PIE models provide interpretation for otherwise puzzling outcomes from various experiments. Although specifics will vary, these principles should apply other niche models involving, say, two energy or immune sources (Fig. 2, 2P2E, 2P2I; see Appendix Section S2). This synthesis of experiments with models underscores need for parametrized trait-based experiments that better predict within-host infection outcomes (Table 1).

Unpacking such within-host mechanisms can improve our understanding of individual health. For instance, deworming trials of hosts coinfected with malaria and gastrointestinal worms show that increased availability of RBCs allow malaria to proliferate within the host, ultimately making them sicker (Budischak *et al* 2018). Disentangling the mechanisms can then allow for correct course of treatment plan to improve individual health (*e.g.,* malarial drugs or vaccines followed by deworming). Together, our work provides a synthesis of within-host niche-based frameworks, laying a theoretical groundwork for a mechanistic understanding of competition outcomes. We then used model predictions to contextualize key findings from the past two decades of experimentation. With this synthesis, we aim to catalyse a new wave of theory-grounded, niche-mechanistic combinations of modelling and experimentation. Ultimately, such a new wave could help to mitigate the severity and comorbidities of coinfections but also advance the development of preventive drug therapeutics and vaccines, ultimately enhancing individual health.

## Data Availability

All data produced in the present study are available upon reasonable request to the authors

## Acknowledgements

AR was supported by the IU Center for the Integrative Study of Animal Behavior, NSF DEB (1655656), and a MSU Presidential Postdoctoral Fellowship. We thank J Lau and the Klausmeier-Litchman lab for helpful comments on the manuscript; D Brisson, J de Roode, S Budischak, S Fellous for generously sharing their data. AR credits J Coltrane and A Ali Khan for artistic inspiration.

